# Utility of family history in disease prediction in the era of polygenic scores

**DOI:** 10.1101/2021.06.25.21259158

**Authors:** Brooke N. Wolford, Ida Surakka, Sarah E. Graham, Jonas B. Nielsen, Wei Zhou, Maiken Elvestad Gabrielsen, Anne Heidi Skogholt, Ben M. Brumpton, Nicholas Douville, Whitney E. Hornsby, Lars G. Fritsche, Michael Boehnke, Seunggeun Lee, Hyun M. Kang, Kristian Hveem, Cristen J. Willer

## Abstract

Clinicians have historically used family history and other risk prediction algorithms to guide patient care and preventive treatment such as statin therapeutics for coronary artery disease. As polygenic scores move towards clinical use, we have begun to consider the interplay of these scores with other predictors for optimal second generation risk prediction. Here, we assess the use of family history and polygenic scores as independent predictors of coronary artery disease and type 2 diabetes. We highlight considerations for use of family history as a predictor of these two diseases after evaluating their effectiveness in the Trøndelag Health Study and the UK Biobank. From these, we advocate for collection of high resolution family history variables in biobanks for future prediction models.

## Perspective

The use of family history in the context of complex diseases may be informative for risk stratification and interventions aimed at prevention. A positive family history of disease puts an individual at a greater than 2 times higher odds of cardiovascular disease^1^ and nearly 3 times greater risk of type 2 diabetes (T2D)^2^. Family history captures inherited genetic variation as well as shared environments and behaviors. Using a statistical framework based on the liability threshold model^3,4^, it was estimated that 32% of the association between parental history and T2D is due to the shared environment between parent-child with the remaining 68% explained by genetics^5^. Family history has been shown to be partially independent from genome-wide polygenic scores (PGSs) in diseases such as schizophrenia^6^ and heart disease^7,8^ despite family history capturing both genetic and environmental disease risk. Other studies have also shown that genome-wide PGSs are associated with incident coronary artery disease (CAD) and T2D are independent of family history^9^.

The simplicity of family history allows for inexpensive and easy to obtain predictive information, potentially allowing for intervention before prolonged exposure to irreversible clinical risk factors, such as smoking or elevated lipid levels. PGSs are more expensive and onerous to obtain than a standard lipid panel or family history, although PGS represents an exposure present from birth that could be ascertained early in life as part of a broad set of risk evaluations. Together, family history and PGSs have the potential to enhance risk prediction in cardiovascular diseases.

### Polygenic scores usher in a new era of risk prediction

Genome-wide association study (GWAS) results are increasingly used to estimate a PGS for individuals by summing over a person’s disease-risk alleles weighted by their impact on disease risk. Studies in CAD shows that individuals with the highest 5% of genome-wide PGSs for CAD have more than a threefold higher risk of CAD than the rest of the population^10^. This is similar to the increased CAD risk conferred by monogenic mutations, such as those causing familial hypercholesterolemia (*LDLR, APOB*, and *PCSK9*). However, 20 times as many people fall into the PGS high-risk category relative to those who carry a monogenic mutation^10^, suggesting that more cardiovascular events could be prevented by selecting individuals based on high PGS in comparison to those with Mendelian mutations. The use of PGS for screening earlier in life is likely preferred to models based on clinical risk factors such as high lipid levels, because individuals falling in the top tail of the PGS distribution typically have earlier disease onset and preventive approaches can be applied prior to development of clinical risk factors. A previous study demonstrated that individuals in the top 2.5% of the PGS distribution were diagnosed with CAD 4.4 years earlier than individuals with average PGS, and for T2D 13.4 years earlier^11^.

### Incorporating family history in an era of polygenic scores

Several studies have evaluated the inclusion of self-reported family history alongside genetics in risk-prediction models for complex diseases such as Crohn’s^12^, CAD^13,14^, breast cancer^15^, and prostate cancer^16,17^. We previously evaluated the use of family history informed genetic risk score (FHiGRS)^18^, and a recently developed method, PRS-FH, combines PGS and family history to improve the accuracy of PGS, particularly in diverse populations^19^. The use of six conventional risk factors for CAD, including family history of heart disease, was shown to improve the prediction of incident CAD when used in combination with PGS compared to prediction based on PGS alone or conventional risk factors alone^20^. Several clinical risk scores (e.g., Reynolds Risk Score, MESA CHD Risk, NORRISK^21^, QRISK^22^) incorporate family history to estimate an individual’s 10-year risk of cardiovascular disease. Family history is formally considered a risk enhancing factor for individuals with an intermediate estimated risk of ASCVD in the United States^23^. However, the Framingham score^24^, SCORE2^25^, and Pooled Cohort Equations (PCE)^26^ do not incorporate family history to estimate 10-year atherosclerotic cardiovascular disease (ASCVD) risk^26^.

Given this background, we examined how existing clinical risk factors such as family history compare to PGS with regards to association with complex disease outcomes. We evaluated prediction using family history and polygenic risk in two independent population-based data sets, the Trøndelag Health Study (HUNT, N=69,635) and the UK Biobank (UKB, N=408,577), for two diseases: CAD and T2D (see Supplemental Methods). We found evidence to support the importance of modeling both family history and PGS for risk prediction in clinical care and observed potentially confounding relationships between self-reported family history and age of the individual at time of self-report. We highlight ways to refine family history and PGS as predictive variables to advance ASCVD risk estimators.

### Proof of concept: family history and PGS as significant predictors of CAD

First, we calculated PGS for CAD (PGS_CAD)_ in 69,635 HUNT study participants based on LDpred and then divided the sample into 20 ventiles, each containing 5% of the sample, to assess the prevalence of disease across the PGS distribution. To assess the impact of family history, we collected self-reported family history reports surveys at the time of study enrollment. We stratified individuals by self-reported family history, divided each stratum into twenty PGS bins (ventiles), and calculated observed CAD prevalence within each family history-stratum and ventile. Notably, as shown in Figure 1, CAD prevalence between strata overlaps only in the opposite tails of PGS distribution—between the top 10% of individuals with no family history of CAD and the bottom 5% of individuals with positive family history of CAD. Since stratification before division into ventiles may bias the results towards larger differences between positive and negative family history strata, we also divided the data by PGS ventiles before stratifying by family history and found the CAD prevalence and trends to be largely similar (Supplementary Figure 1). In a sensitivity analysis across the number of quantile divisions (from 4 to 100), the trend between negative and positive family history strata was robust (Supplementary Figure 1).

**Figure 1:**
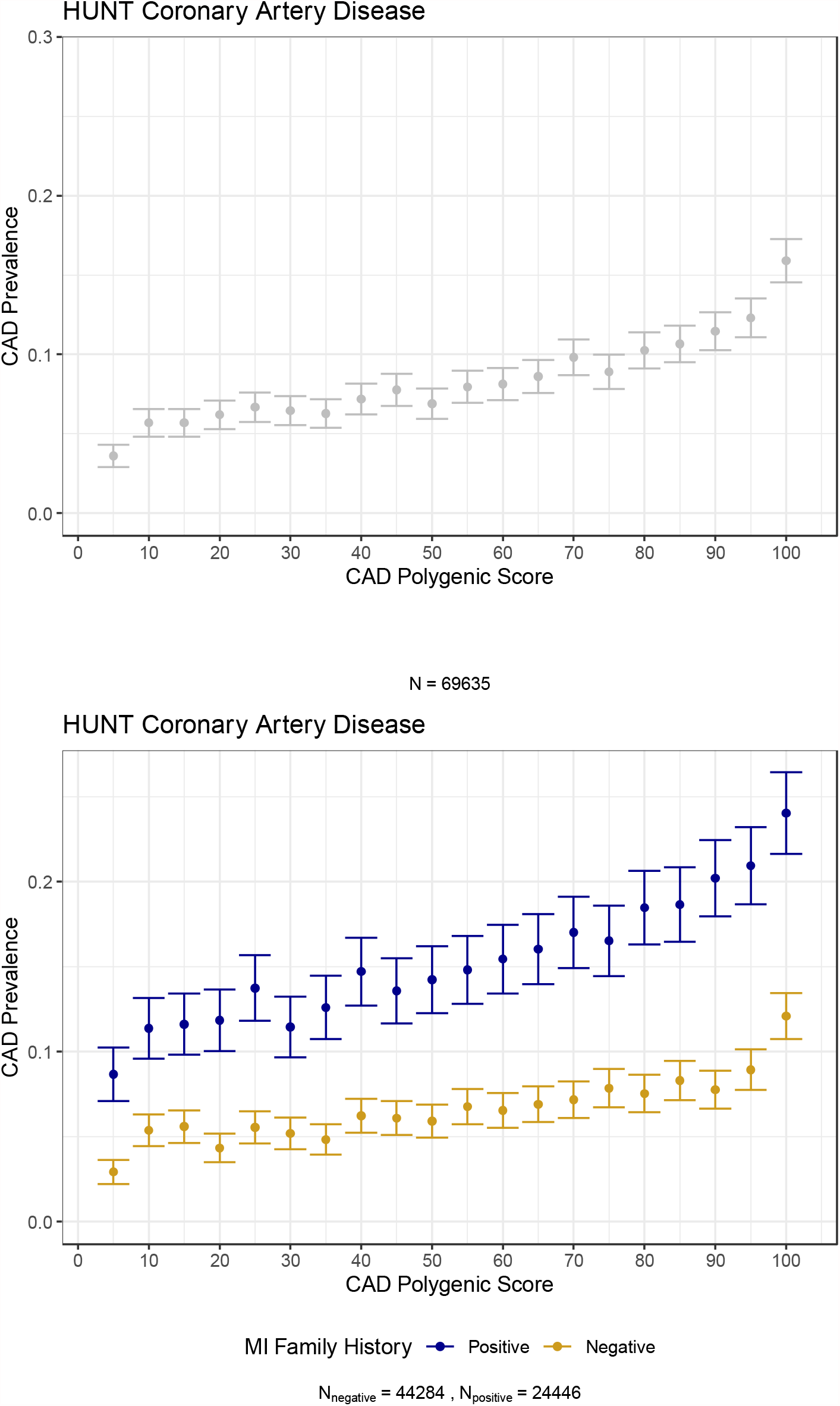
CAD prevalence across PGS quantiles, stratified by family history of myocardial infarction in HUNT. Prevalence of coronary artery disease per polygenic score ventile in the entire population of HUNT, stratified by self-reported family history of myocardial infarction (MI).

In HUNT participants with a positive family history of CAD, individuals with a CAD polygenic score (PGS_CAD_) in the top 5% of the score distribution had 2.78 times higher odds of CAD (95% CI 2.41-3.22) compared to 2.59 times higher odds of CAD among all participants with high PGS_CAD_ (95% CI 2.34-2.87) (Table 1). This trend—of larger odds of disease in the high-risk group stratified first by family history and then by PGS_CAD_ —holds across quantile thresholds for top scores (e.g. 5%, 10%, 20%, Table 1).

**Table 1.**
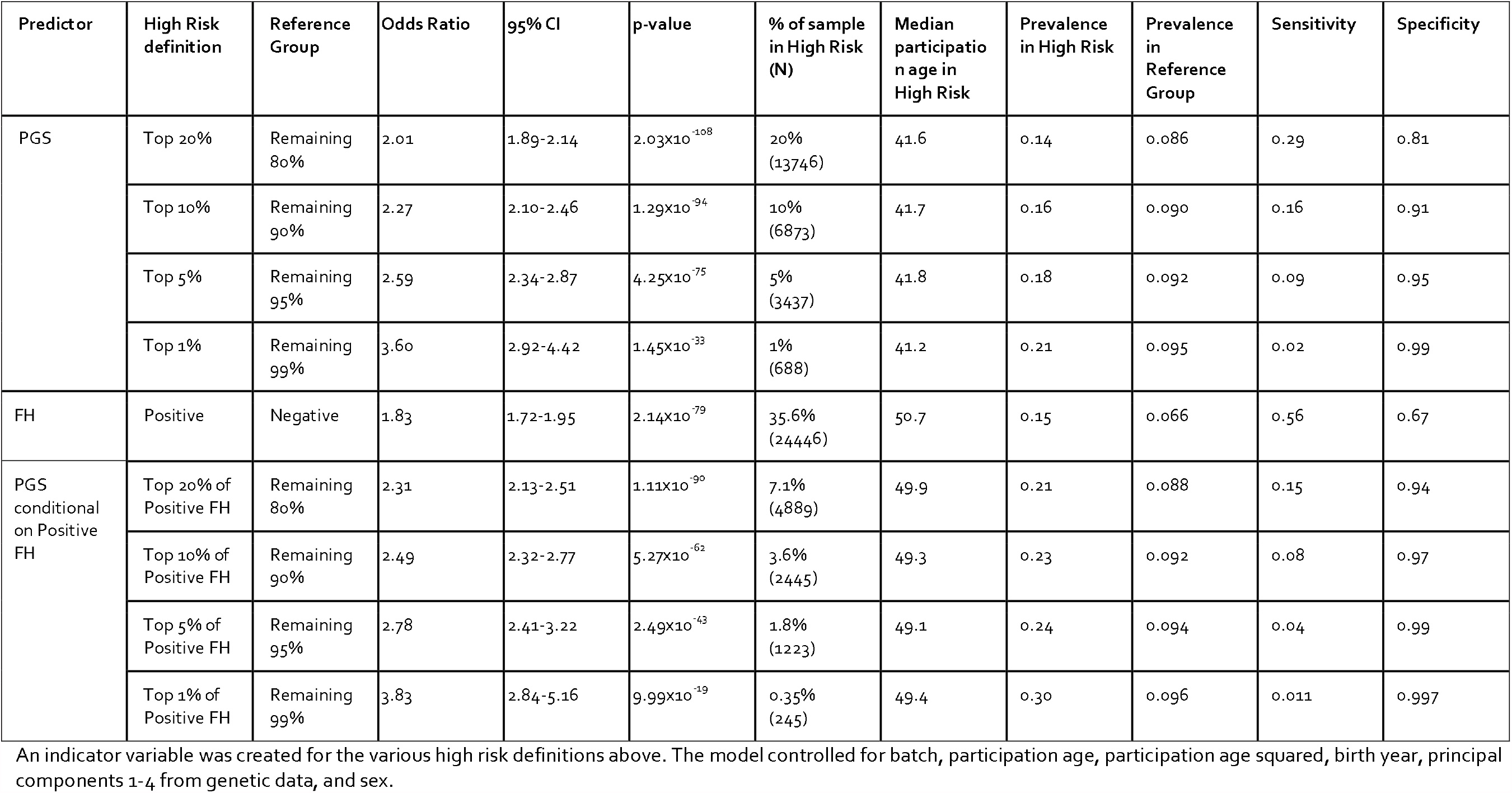
Clinical impact of high risk stratification for CAD in HUNT.

The PGS_CAD_ distributions are significantly different between CAD cases and controls (Wilcoxon Rank Sum Test [WRST] p-value=1.4×10^−127^), and between positive and negative self-reported family history (WRST p-value=1.5×10^−125^, Supplementary Figure 2). The Pearson correlation between PGS_CAD_ and family history is 0.09 (Supplementary Figure 3). While this correlation is low, we observed a significant association between family history and PGS_CAD_using a logistic regression model (p-value=4×10^−131^, OR for positive family history=1.22 per s.d. of PGS_CAD_ [1.20,1.24]).

Through model selection, we observed that birth year and participation age (also known as biobank enrollment age) were significant predictors for CAD. Using a full model, we demonstrate that family history and PGS_CAD_ are significant predictors of disease (Table 2), even after accounting for birth year and enrollment age, with a high degree of independent information. A positive family history puts an individual at nearly 2 times greater odds of CAD (OR=1.72, 95% CI 1.61-1.83, Table 2). Family history and PGS_CAD_ have a nominally significant interaction term (p-value=0.02) in the full model (Table 3). Adding PGS_CAD_ to the base model yields a larger change in Nagelkerke’s R^2^ (0.023) than adding family history to the base model (0.010) (Table 3).

**Table 2.** Full model estimates for CAD in HUNT.

| Predictor | OR | 95% CI | p-value |
| --- | --- | --- | --- |
| Standardized Participation Age | 10.9 | 8.5-14.0 | $2.96 \times 10^{-76}$ |
| Standardized Participation Age Squared | 0.13 | 0.11-0.16 | $1.21 \times 10^{-86}$ |
| Standardized 2021-birthYear | 2.86 | 2.62-3.10 | $1.94 \times 10^{-135}$ |
| Male Sex | 2.69 | 2.54-2.85 | $4.25 \times 10^{-253}$ |
| Positive Family History | 1.72 | 1.61-1.83 | $3.39 \times 10^{-60}$ |
| Inverse normalized PGS | 1.53 | 1.53-1.60 | $3.66 \times 10^{-9}$ |
| Family History x Inverse normalized PGS (interaction term) | 0.94 | 0.86-0.99 | .024 |

Adjusted for principal components 1-4 from genetic data and genotyping batch (HUNT)

**Table 3.** Model comparisons for CAD in HUNT.

| Model 1 | Model 2 | LRT p-value | ⊗ Nagelkerke's $r^2$ |
| --- | --- | --- | --- |
| Base | PGS model | $9.22 \times 10^{-188}$ | 0.023 |
| Base | FH model | $8.72 \times 10^{-82}$ | 0.010 |
| PGS model | PGS + FH (additive) model | $1.71 \times 10^{-60}$ | 0.007 |
| FH model | PGS + FH (additive) model | $1.65 \times 10^{-166}$ | 0.021 |
| PGS + FH (additive) model | PGS + FH + PGS x FH (interaction) model | 0.022 | 0.00014 |

Comparison of models in HUNT with family history (FH) and polygenic score (PGS) using ANOVA. The base model is sex, birthyear, participant age, and participant age squared, and first four principal components from genetic data

### Age matters with respect to family history in risk prediction models

Older individuals are more likely to report a positive family history (Figure 2). The Pearson correlation between enrollment age and positive family history of myocardial infarction (MI) was 0.38 (Supplementary Figure 3). This has important implications for: i) recording age at variable collection, ii) the importance of updating family history within the electronic medical record, iii) determination of optimal age to use family history in risk prediction, and iv) the impact of pharmaceutical intervention on disease and family history incidence.

**Figure 2.**
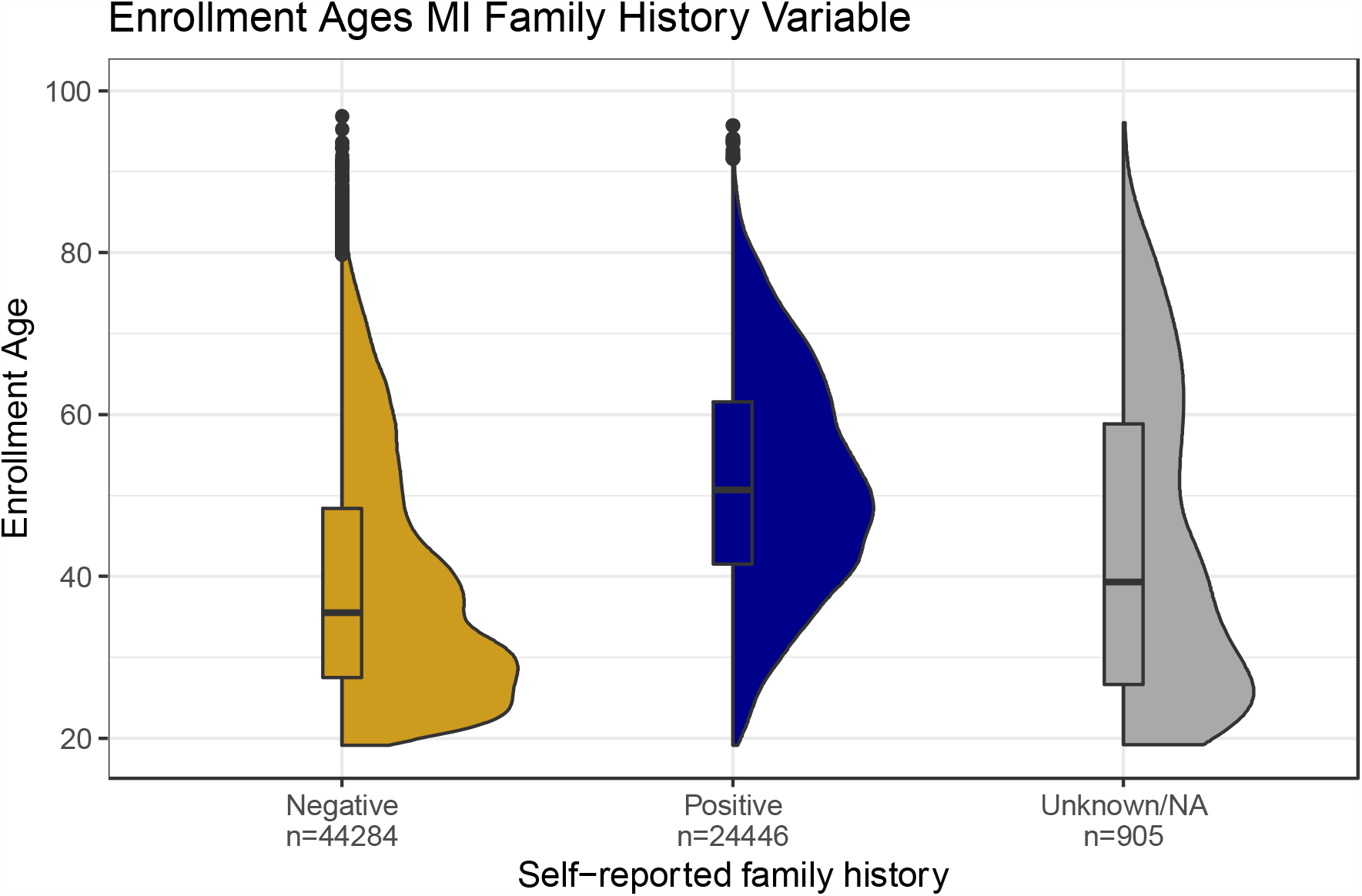
Distribution of participation ages (i.e. biobank enrollment ages) for the first degree family history of myocardial infarction.

The average age of first MI in HUNT was 70.5 years (95% CI 70.3,70.9). A positive family history for MI was significantly predicted by older age at enrollment (2-sided p-value < 2 × 10^−308^). HUNT2 participants were asked if they have a family member who had an MI before the age of 60: sixteen percent of participants between 19-40 years of age reported “yes” versus 52% of participants over 40 years of age. Similarly, the median enrollment age of persons reporting no affected first degree relative was significantly lower than the age of persons reporting positive family history (35.5 versus 50.7 years, WRST 1-sided p-value < 2.2 × 10^−308^, Figure 2). In HUNT2, the survey metric specified the relationship type experiencing a MI before 60 years of age. Individuals that reported a sibling or child with the disease were slightly older than individuals who reported affected parents (48.7 versus 48.2 years, WRST 1-sided p-value=7.9×10^−11^).

### Enrollment Ages MI Family History Variable

This finding is not surprising for common, complex diseases—as someone ages, their relatives also age and are at a higher risk of disease. However, this finding encourages careful collection of the age at which an individual self-reports family history of disease. Presently, the accuracy of self-reported family history is imperfect, with some studies indicating specificity ranging from 75-98% for common conditions such as diabetes and obesity^27^. We found age at self-report of family history was an important variable to control for when using family history in prediction models (Supplementary Figure 4). Within the HUNT longitudinal study, we identified instances where more recent family history data were used to correct or update past family history variables from questionnaires, which de-coupled family history from the reporting age. As such, the individual’s age at time of self-reporting family history was incorrect for a small subset of individuals whose family history record had been updated.

We tested the performance of the predictors—family history and PGS_CAD_—with respect to age at self-report by modeling across enrollment age bins (e.g., the age an individual was when they completed the questionnaire and self-reported a positive or negative family history). Both family history and PGS_CAD_ were significant predictors across the lifespan for CAD (Figure 3). Family history of MI showed a U-shaped curve and had a maximum odds ratio estimate at the youngest enrollment age bin (19-30). We hypothesize the high effect of family history between enrollment ages 19-30 is driven by rare variants of large effect, leading to earlier onset or more severe disease. The higher odds ratio observed for older enrollment ages for family history may be due to lifetime exposure to shared-family environmental risk factors (e.g., diet, exercise, smoking) and more time for cardiac events driven by polygenic genetic risk to occur in family members. The odds ratio estimate for PGS_CAD_ decreases slightly across decades for CAD. We hypothesize that environmental factors introduce more variation into the outcome as a person ages, so the contribution of genetics to risk decreases concomitant with an increase in the role of environmental and lifestyle risk factors. In comparison to family history, the predictive utility of PGSs were much more consistent across age of study participant.

**Figure 3.**
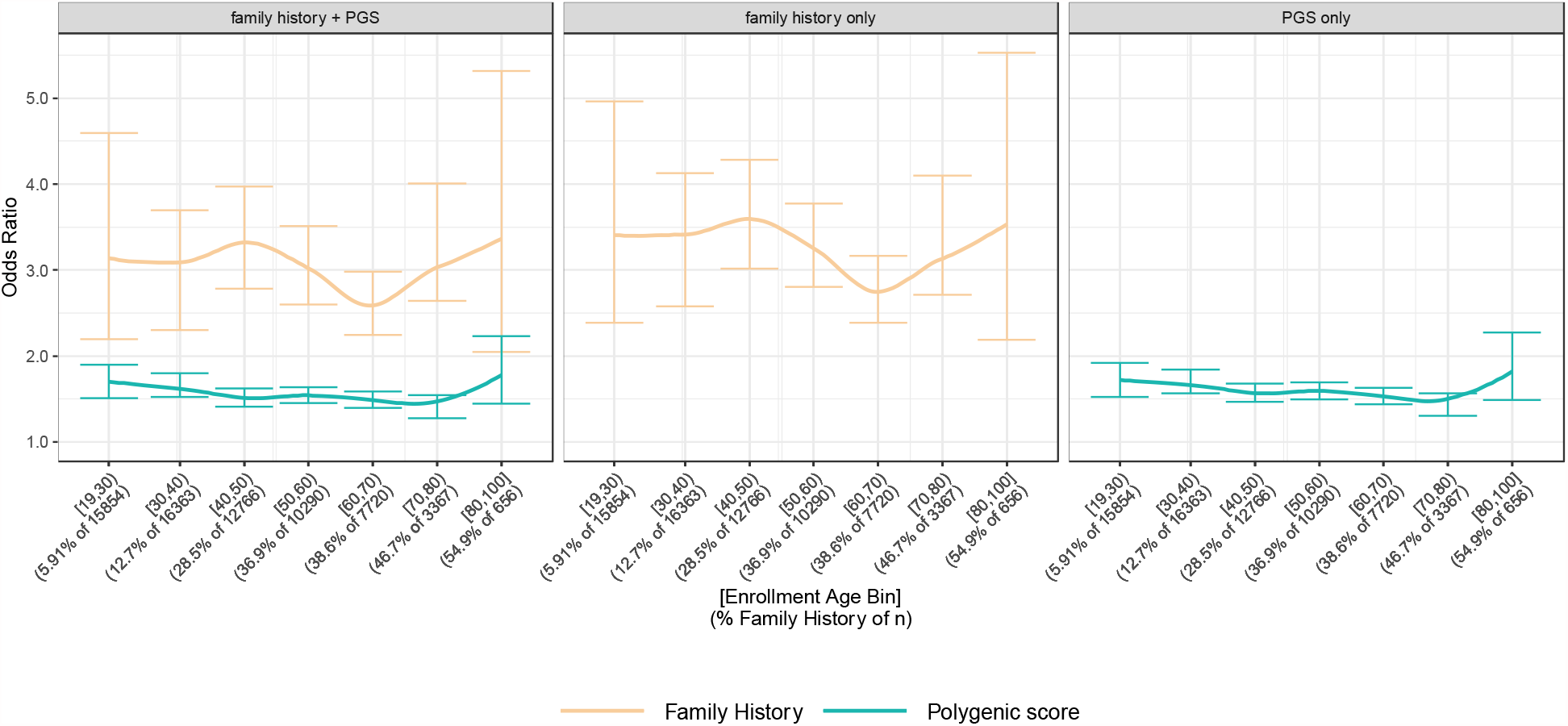
Family history and PGS as predictors of CAD across biobank enrollment ages. Each model is adjusted for principal components 1-4 from genetic data, participation age, participation age squared, birthyear, sex, and genotyping batch Odds ratio for PGS is for continuous PGS (yellow) and for FH is for positive family history (green) within the enrollment age bin and for.

At first glance, family history is an ideal predictive indicator for CAD, since it is inexpensive and easy to obtain, however, the paucity of familial disease events for young individuals (Figure 2) suggests family history may be a less effective predictive tool than PGS for early intervention. Before 40 years of age, we expect that family history might help capture individuals at risk due to familial monogenic mutations causing early onset disease in parents, but is probably not as helpful in cases of polygenic genetic variation causing later disease onset (Figure 3). By the time a sibling is old enough to become affected, the benefit of family history as a disease predictor is less useful as the timeframe for preventive interventions for the individual may have mostly passed. A tool that has its greatest predictive effect after the average age of disease onset is likely less effective. This finding may prove to limit the utility of family history to predict late-onset diseases, particularly diseases observed in siblings or cousins.

As more effective preventive strategies are introduced and rates of cardiovascular disease decrease in the population, we expect rates of positive family history to decrease in frequency. While this will be a welcome outcome of precision medicine, it does have ramifications for predictors such as family history which are a function of disease incidence. This has been observed for individuals with familial hypercholesterolemia, in whom high-intensity lipid-lowering therapies have dramatically decreased the risk of MI^28^. As of 2013, 27.8% of the general adult (>40 years of age) population in the United States report using statins, and 52.7% of patients with ASCVD use statins^29^. Recent research suggests high-intensity statin usage could prevent 51-71% of premature ASCVD events (1.4 million events in the US) when patients aged 30-39 are treated for 30 years^30^. Using genetically inferred kinship in the subset of HUNT for which we have statin information (HUNT3, N=14,055), of the 2,595 first degree relatives of cases, 26.8% take statins compared to 16.8% of individuals not related to a case (Chi-square p-value=3.6×10^−58^). A person with a high risk of ASCVD may have relatives on statins, which prevents disease progression, and therefore report a negative family history. For this reason, the utility of family history as a predictor across the lifespan will need detailed evaluation and may change for different generations.

### Family history and PGS trends replicate for Type 2 Diabetes

We evaluated the same models for T2D, another complex disease with environmental and genetic risk factors and with well-powered GWAS available for the PGS. Similar to CAD, we observed an overlap of the family history strata with the top 5% of PGS_T2D_ individuals with no family history of T2D and the bottom 5% of PGS_T2D_ individuals with positive family history (Figure 4). Participants with a PGS_T2D_ in the top 5% with a positive family history have 3.64 times higher odds of T2D compared to the rest of the population, versus 2.6 times higher odds without stratification by family history (Supplementary Table 1). The PGS_T2D_ distributions are significantly different between T2D cases and controls (WRST p-value=3.3×10^−173^) and between positive and negative self-reported family history (WRST p-value=3.4×10^−96^). While the Pearson correlation between family history and PGS_T2D_ is small (r=0.08, Supplementary Figure 5), the association between T2D and PGS_T2D_ was significant (p-value=3×10^−8^, OR=1.21 [1.19,1.24]). A positive family history was associated with 3 times greater odds of having T2D (OR=3.01, 95% CI 2.79-3.24, Supplementary Table 2). We observed a larger increase of Nagelkerke’s R^2^ when adding family history to PGS_T2D_ with T2D compared to CAD (0.026 versus 0.021, Supplementary Table 3). Family history has a larger association with T2D than CAD, potentially because it represents more of a shared environmental component, or because there is not as substantial a depletion of positive family history for T2D due to drug treatment as there is for CAD due to treatment with statins.

**Figure 4.**
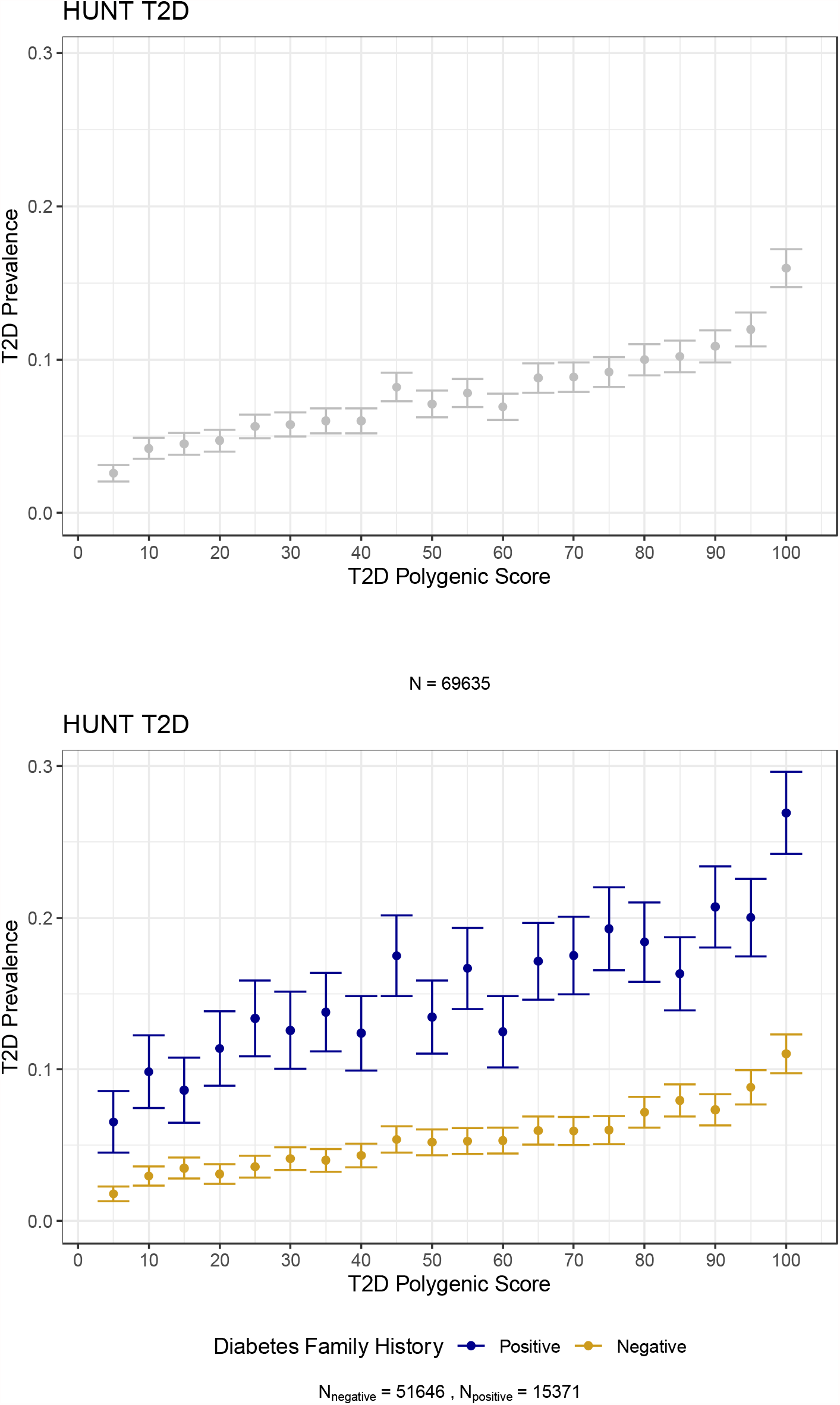
T2D prevalence across PGS quantiles, stratified by family history of diabetes in HUNT. The prevalence of Type 2 diabetes per polygenic score ventile in the entire population of HUNT and stratified by self-reported family history of diabetes.

Similar to the observations for CAD, the Pearson correlation between age of enrollment and family history of T2D is 0.33 (Supplementary Figure 5). Nine percent of 19-40 year aged participants report a positive family history of T2D, versus 35% of participants greater than 40 years of age. Both PGS and family history (when modeled together) are significant across the lifespan for T2D (Figure 5). The odds ratio estimated for family history of T2D had a U-shaped curve with higher odds of disease indicated by family history on both tails of enrollment age (Figure 5), again similar to the pattern observed for CAD association with family history of MI.

**Figure 5.**
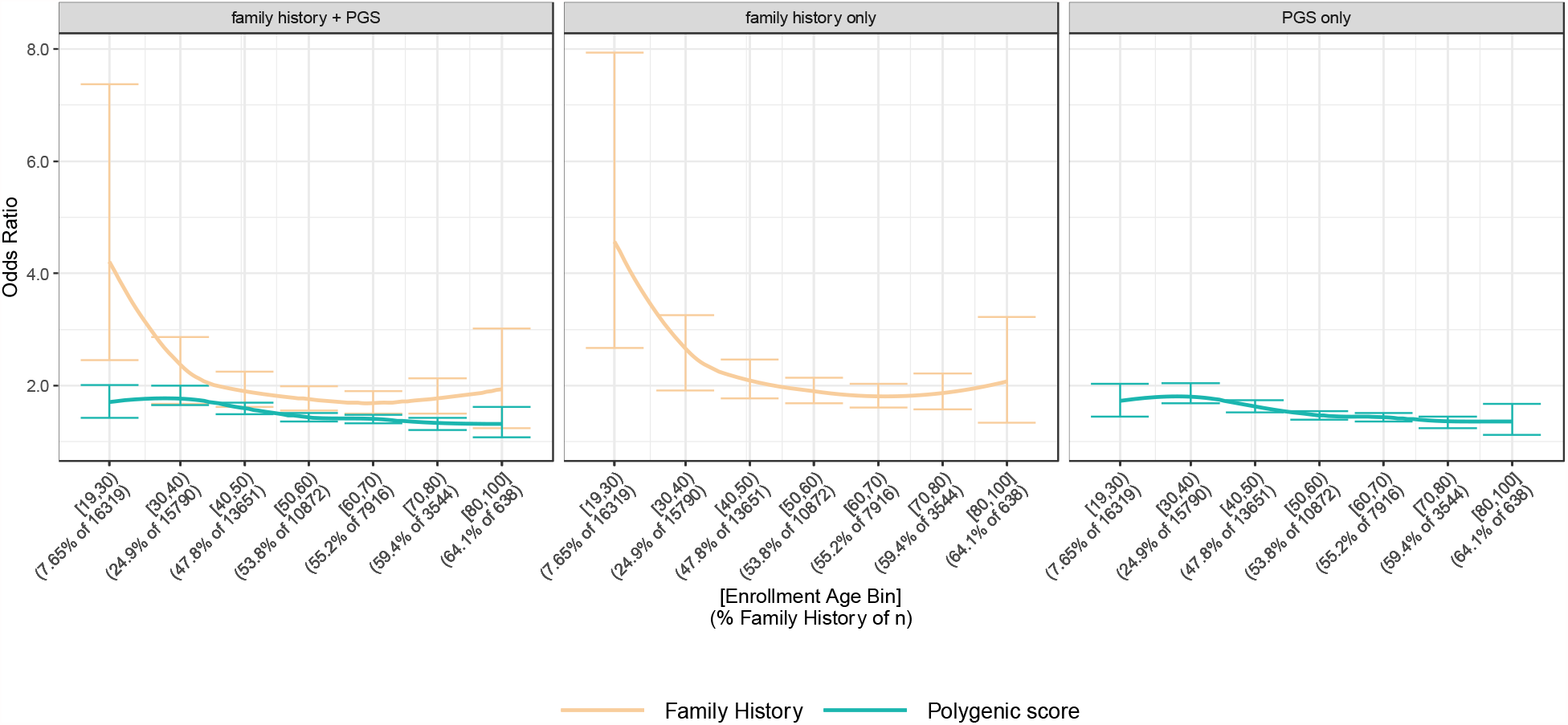
Family history and PGS as predictors of T2D across biobank enrollment ages in HUNT. Each model is adjusted for principal components 1-4 from genetic data, participation age, participation age squared, birthyear, sex, and genotyping batch.

### Family history and PGS trends replicate in the UK Biobank

When we assessed the relationships between family history, polygenic score and disease prevalence in the UK Biobank, an increased disease prevalence was observed in individuals in the top tail of the PGS distribution with a positive self-reported family history for both CAD (Supplementary Figure 6) and T2D (Supplementary Figure 7). A relationship between negative family history for heart disease and younger enrollment ages was also observed (Supplementary Figure 8). Using the covariates from the model selection from HUNT, we observed similar odds ratios for clinical predictors in UK Biobank as in HUNT (Supplementary Table 2,4). In the UK Biobank, a model with predictors for both family history and PGS and their interaction were significant terms for CAD, but the interaction term between PGS and family history was not significant for T2D (Supplementary Table 5).

### Improving family history and polygenic scores to advance prediction of CAD

Current American Heart Association guidelines for lipid-lowering (i.e., statin, ezetimibe or PCSK9i therapies) intervention are multifaceted with a many-step protocol based on: past CVD events, LDL-C levels, 10-year ASCVD risk estimated by PCE, diabetes status, age, and coronary artery calcium score^23^. Family history is considered a risk enhancing factor, however, we advocate for formal inclusion of both family history and PGS to risk estimation models given that both variables are significant and independent predictors of CAD in HUNT and UK Biobank. Currently PGSs are limited by trans-ethnic portability^31,32^, sensitivity to population stratification^33^, and miscalibration^34^ among other considerations. Future iterations of PGSs may integrate genetic risk for clinical risk factors such as genetic prediction of LDL cholesterol or body mass index or other multi-trait risk models that improve prediction. The addition of an easily ascertained predictor such as family history suggests we should incorporate this variable as we continue to evaluate the use of other biomarker PGSs (as in Sinnott-Armstrong *et al*^35^) and clinical risk factors to predict disease (as in Inouye *et al*^20^), particularly early in life.

Family history must be consistently recorded in the electronic health record to be impactful in advanced risk estimation algorithms. For example, a binary predictor describing the presence or absence of family history is less informative than more precise family history records such as: age at time of family history report, the number of affected relatives, relationship to relatives with disease, severity of disease in the family member, or the age of disease onset/diagnosis in these relatives. Differentiating between first-degree relative (mother, father, sibling) and second degree relative (grandparent, aunt, uncle) will yield specificity as to the degree of shared genetic liability. Even more useful is a grid of diseases and relationships to allow for higher resolution family history variables. As providers move towards electronic surveys at intake of clinical appointments, logic allowing for more detailed questions about family members with specific diseases listed on the grid should be implemented. The age at time of reporting family history should be recorded and regular updates to both the family history information (coupled with age at time of report) will improve prediction based on family history.

These richer predictive features are rarely systematically collected in biobank surveys, clinic visits, or the electronic health record, and we contend that this detailed documentation will enable greater predictive accuracy and contribute to earlier intervention with preventive therapies. We observed similar levels of utility and independence of family history and PGS association with T2D as with CAD. In the absence of quantitative risk prediction algorithms for T2D (such as the PCE for CAD), our work suggests the potential utility of family history and PGS in addition to clinical measurements such as HbA1C. Additional studies should be performed in traits with Mendelian inheritance patterns (e.g., breast cancer) and early onset diseases (e.g., asthma) to determine the utility of family history across a spectrum of disease prevalence, heritability and genetic architecture.

## Conclusion

In two electronic health record-linked biobanks, HUNT and UK Biobank, we evaluated the association of family history and PGS with two different diseases: CAD and T2D. We confirm that family history and PGS are both significant and mostly independent predictors of disease by evaluating CAD and T2D prevalence. Given the significant but weak interaction between family history and PGS, we note that family history is not simply a proxy for PGS, but likely represents lifestyle and social determinants of health, and is therefore, an important component of risk prediction in addition to PGS. We demonstrate increasing rates of positive family history with increasing age at report of family history. We also highlight that positive family history of MI is less common at younger ages, when relatives are also young, but family history also has the highest impact on odds of CAD in this age group. We suggest advancing electronic health record-linked biobank infrastructure to enable meaningful integration of detailed family history and PGS to improve upon current ASCVD risk estimation with PCE leading to prevention of disease.

## Supporting information

SuppMaterials

## Data Availability

Code is publicly available on GitHub (https://github.com/bnwolford/FHiGR_score) and summary statistics are made available in the manuscript. Individual level data is not available for participant privacy.

## Acknowledgements

We thank all research participants in the HUNT study and the UK Biobank for their dedication towards improving human health. We thank Bethany Klunder for project management. This research has been conducted using the UK Biobank Resource under application number 24460. The HUNT-MI study, which comprises the genetic investigations of the HUNT Study, is a collaboration between investigators from the HUNT study and University of Michigan Medical School and the University of Michigan School of Public Health. The K.G. Jebsen Center for Genetic Epidemiology is financed by Stiftelsen Kristian Gerhard Jebsen; Faculty of Medicine and Health Sciences, NTNU, Norwegian University of Science and Technology (NTNU) and Central Norway Regional Health Authority. The Trøndelag Health Study (HUNT) is a collaboration between HUNT Research Centre (Faculty of Medicine and Health Sciences, Norwegian University of Science and Technology NTNU), Trøndelag County Council, Central Norway Regional Health Authority, and the Norwegian Institute of Public Health.

## URLs

https://github.com/bnwolford/FHiGR_score.

## Conflict of Interest

C.J.W.’s spouse works for Regeneron Pharmaceuticals. J.B.N. is employed by Regeneron Pharmaceuticals, Inc.

## Funding

Funding for this study was provided by the National Institutes of Health to C.J.W. (R01 HL127564, R35 HL135824) and M.B. (U01 DK062370, R01 HG009976). B.N.W. was funded by NSF Graduate Research Fellowship (DGE 1256260) and NIH Training Program in Genomic Science (T32 HG000040). N.J.D. was funded by a Foundation for Anesthesia Education and Research (FAER) Mentored Research Training Grant. W.Z. was supported by the National Human Genome Research Institute of the National Institutes of Health under award number T32HG010464.

## Author Contributions

B.N.W. and C.J.W. designed the study. B.N.W. performed most of the primary analyses, with assistance from I.S. B.N.W. wrote the manuscript and I.S., C.J.W., and W.E.H. revised. All other authors contributed to study implementation.

## Supplementary Methods

### Trøndelag Health Study

The Trøndelag Health Study (HUNT) is a population-based health survey conducted in Trøndelag county, Norway, since 1984^36^. Participation in the HUNT Study is based on informed consent, and the study has been approved by the Data Inspectorate and the Regional Ethics Committee for Medical Research in Norway. Of the >120,000 participants in the HUNT 1-3 study, 69,635 individuals of European ancestry have been genotyped using Illumina Human CoreExome v1.1 array with 70,000 additional custom content beads and imputed to 25M genetic markers using 2,202 whole-genome sequenced samples from HUNT together with Haplotype Reference Consortium reference panel^37,38^. We used a combination of hospital, outpatient, and emergency room discharge diagnoses (ICD-9 and ICD-10) along with self-reported variables and lab measurements to identify cases and controls for common diseases (Supplementary Table 6,7). Self-reported family history of disease was obtained from survey questionnaires from HUNT 1-3 (Supplementary Table 8). Variables across HUNT collections were collapsed to create a single indicator variable for first-degree family history of myocardial infarction (MI) or diabetes for as many samples as possible. The age of participation in HUNT 1-3 was recorded with the earliest age being taken if the participant answered the question in multiple collections.

### UK Biobank

The UK Biobank is a population-based cohort collected from multiple sites across the United Kingdom^39,40^. Genotyped and imputed data for 408,577 individuals of white British ancestry were used for this analysis. Case and control status was ascertained using phecodes^41^. Family history across multiple family members was obtained from field IDs 20107, 20110, 20111 and collapsed into a single indicator variable for first degree family history of heart disease or diabetes (Supplementary Table 6-8).

### Polygenic scores

We used previously generated weights for an optimized set of genome-wide variants (6.6M for CAD and 6.9M for T2D) to calculate the disease-specific PGS^10^. Briefly, these weights^6^ were based on genetic effect estimates (beta) from large GWAS for CAD (N=60,801 cases and 123,504 controls) and T2D (N=26,676 cases and 132,532 controls) and genetic variants were pruned using LDpred and tuning parameters of 0.001 and 0.01 respectively. The weights for CAD and T2D were applied to individual-level imputed dosages for each HUNT participant and UKB participant to estimate PGS_CAD_ and PGS_T2D_. A limitation of this analysis is the LDpred tuning parameters were optimized in UKB phase 1, but the weights came from external GWAS and the performance did not vary widely across the models in the optimization step.

### Statistical analysis

We estimated the odds ratios (ORs) for models with PGS and self-reported family history as predictors using logistic regression with binomial link function adjusting for covariates including the effect of sex, age at biobank enrollment, age at biobank enrollment squared, birth year, and first four genetic principal components. In analyses where we estimate the odds ratio for predictors, we perform several variable transformations. Birth year is transformed to the age in 2021 so the odds ratio is on the scale of risk rather than protection (i.e odds ratio > 1), but is referred to as birth year to avoid confusion. The PGS is inverse normalized (using R package RNOMni) and age-related covariates are scaled to have a mean of 0 and variance of 1. When evaluating model selection for family history and PGS we used standard multivariable logistic regression. When considering risk thresholds using family history and PGS, we used an indicator variable based on a percentile threshold for PGS with or without conditioning on family history. Reported p-values from logistic regression are from Wald tests, and the p-values from model comparison with ANOVA are Likelihood Ratio Tests. Statistical analyses were conducted using R version 4.0.3 software. Hereafter, when describing the predictors, family history refers to self-reported family history from surveys

## Notes

### Author Declarations

The HUNT study has previously received human subjects approval at the University of Michigan and the Norwegian University for Science and Technology (NTNU). The Institutional Review Boards of the University of Michigan Medical School (IRBMED) provided ethical approval under HUM00126227. The Regionale komiteer for medisinsk og helsefaglig forskningsetikk (REK) provided ethical approval with REK-midt approving study 2015/1209 and REK-sø r-øst approving study 2016/885. All study participants provided written informed consent and are consented for dbGaP deposition. HUNT subjects were recruited from a region of Northern Norway, and the total number of participants in the HUNT Biobank is 78,951 individuals. We have obtained human subjects approval for sequencing studies on HUNT subjects both from NTNU and University of Michigan. Sharing of de-identified materials is covered by existing informed consents and IRB.

