## Supplementary material for "Utility of family history in disease prediction in the era of polygenic scores": SuppMaterials

### Supplementary Figures and Tables

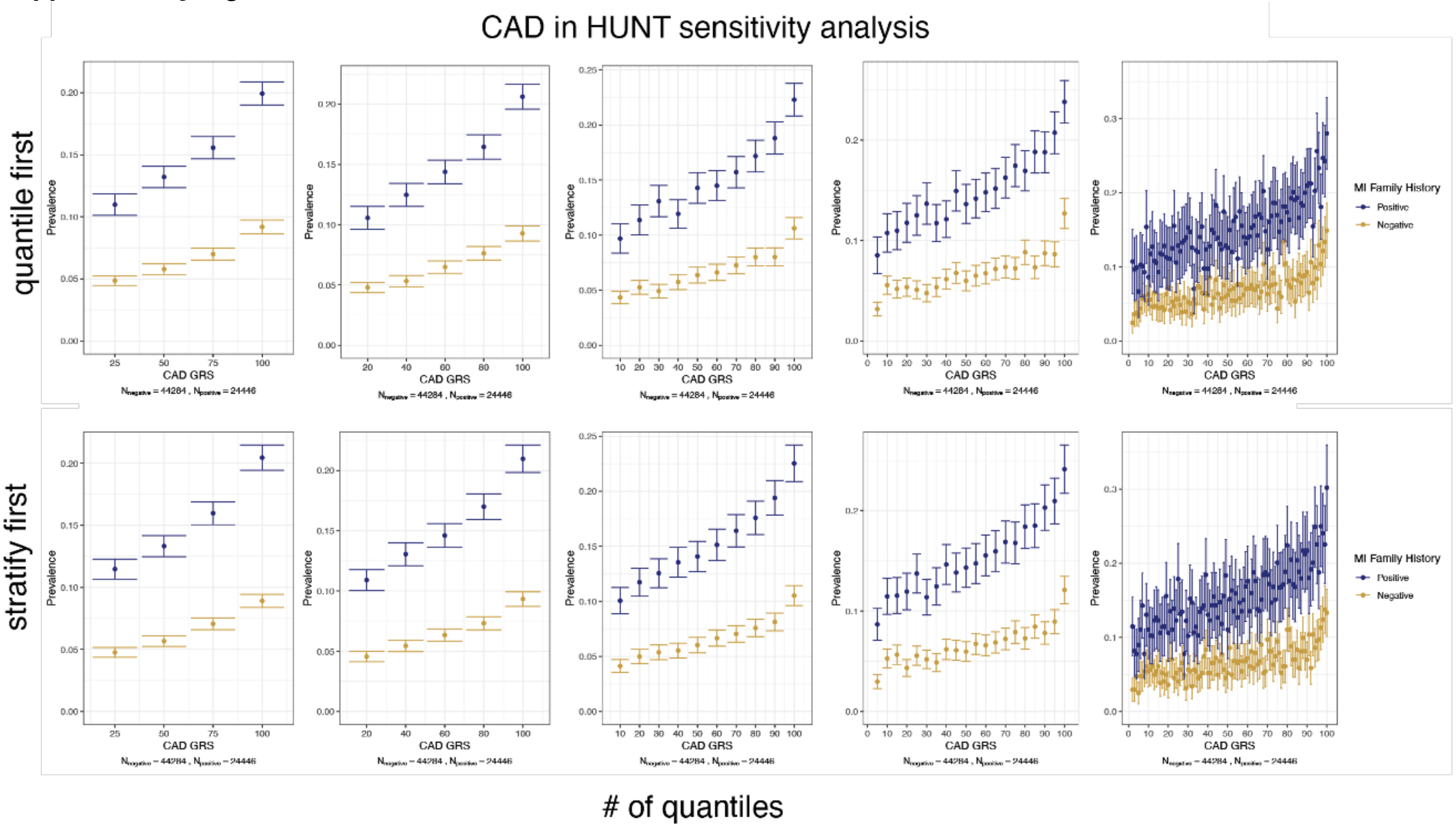


**Supplementary Figure 1 Sensitivity analysis for disease prevalence**

Regardless of the number of quantiles (n=4,5,10,20,100) or if quantiles are calculated before (quantile first) or after (stratify first) stratification by family history, the trends remain.

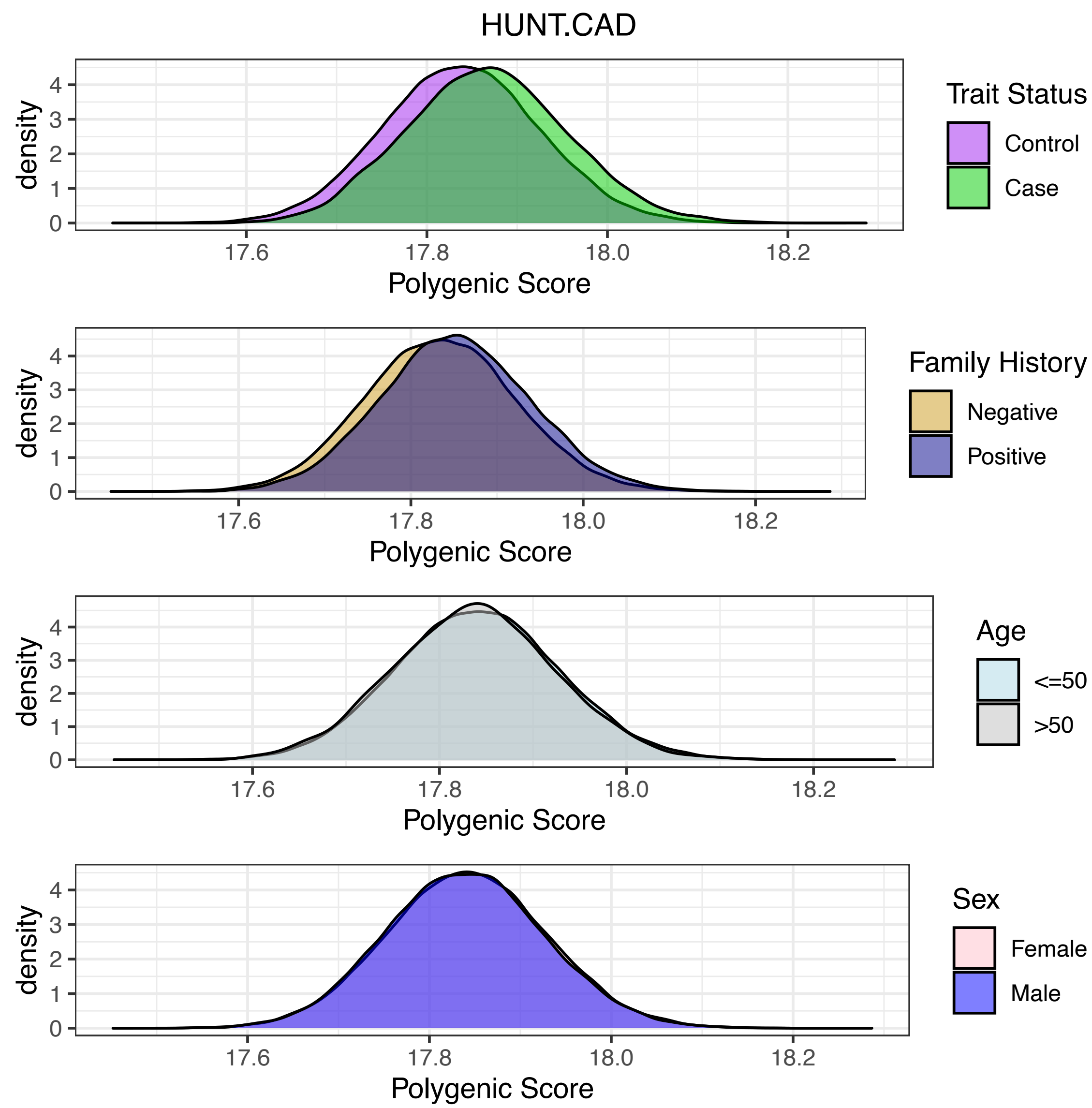


**Supplementary Figure 2 Distribution of PGS for CAD in HUNT**

Inverse normalized polygenic scre stratified by trait status, family history, 50 years of age, and sex.

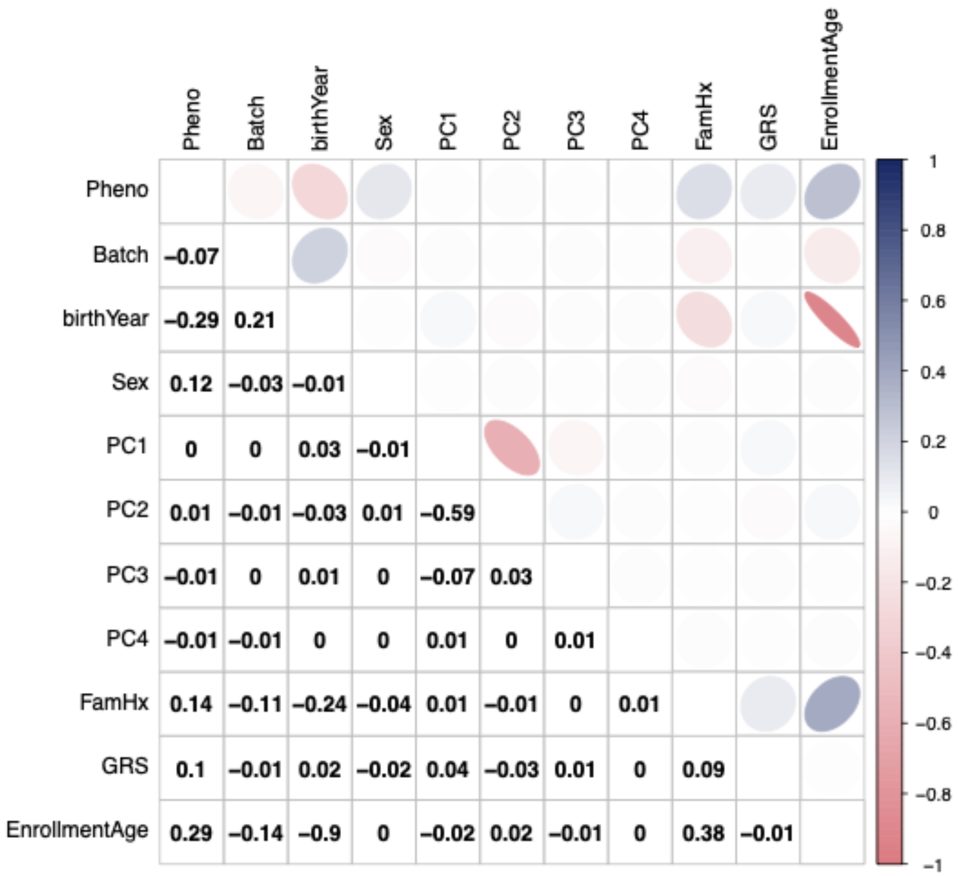


**Supplementary Figure 3 Pearson correlations between model variables for CAD in HUNT.** Pheno is the phenotype (e.g., CAD). Batch is genotyping batch coded 0,1. FamHx is family history coded 0,1. Sex is coded 0= females and 1= males. Enrollment age is the age at which a participant filled out the self-report family history variables in a HUNT survey.

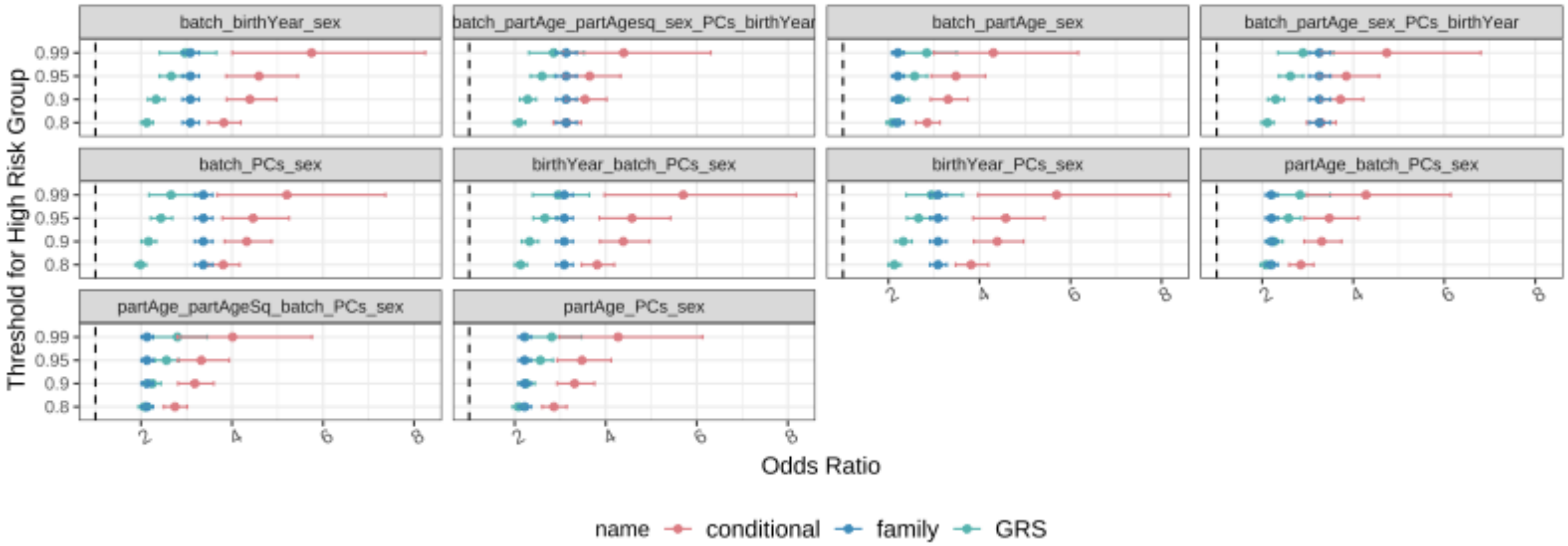


**Supplementary Figure 4 Model selection for CAD**

An indicator variable was used to identify a "high risk" group. Conditional is top X% of distribution with positive family history. Model selection was performed, leaving out one covariate at a time. Batch is genotyping batch, participation age is the age family history was self reported, partAgesq is participation age squared. All continuous variables were scaled to mean of 0 and variance of 1. PGS was inverse normalized. When birthyear and participation age are not included, family history has a higher odds ratio than when these covariates are adjusted for

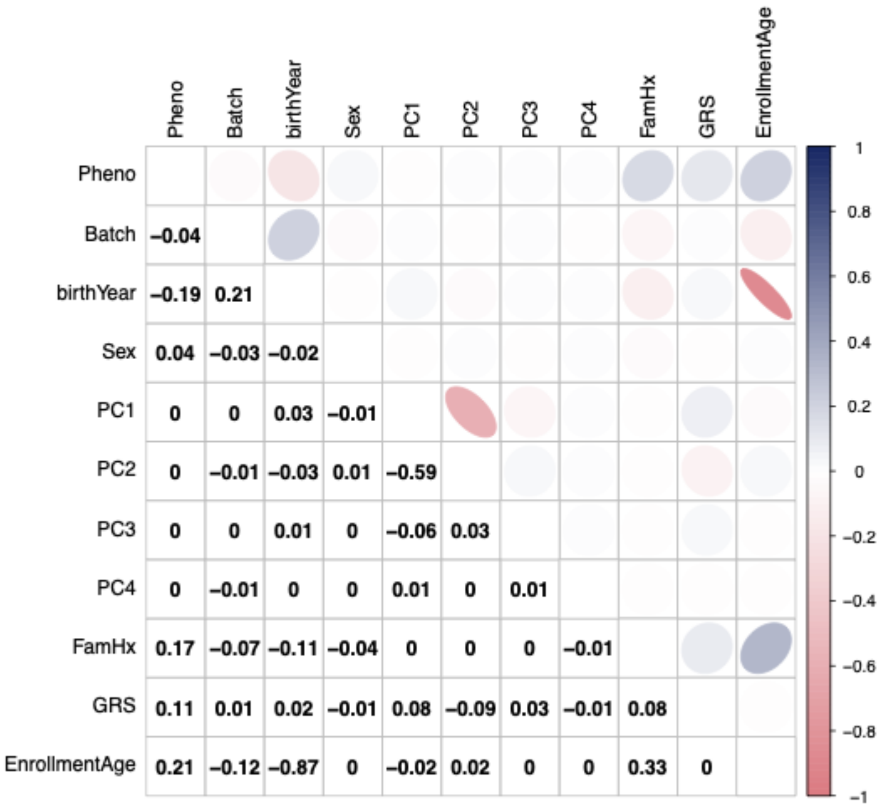


**Supplementary Figure 5 Pearson correlations between model variables for T2D in HUNT.**

Pheno is the phenotype (e.g., Type 2 diabetes). Batch is genotyping batch coded 0,1. FamHx is family history coded 0,1. Sex is coded 0= females and 1= males. Enrollment age is the age at which a participant filled out the self-report family history variables in a HUNT survey.

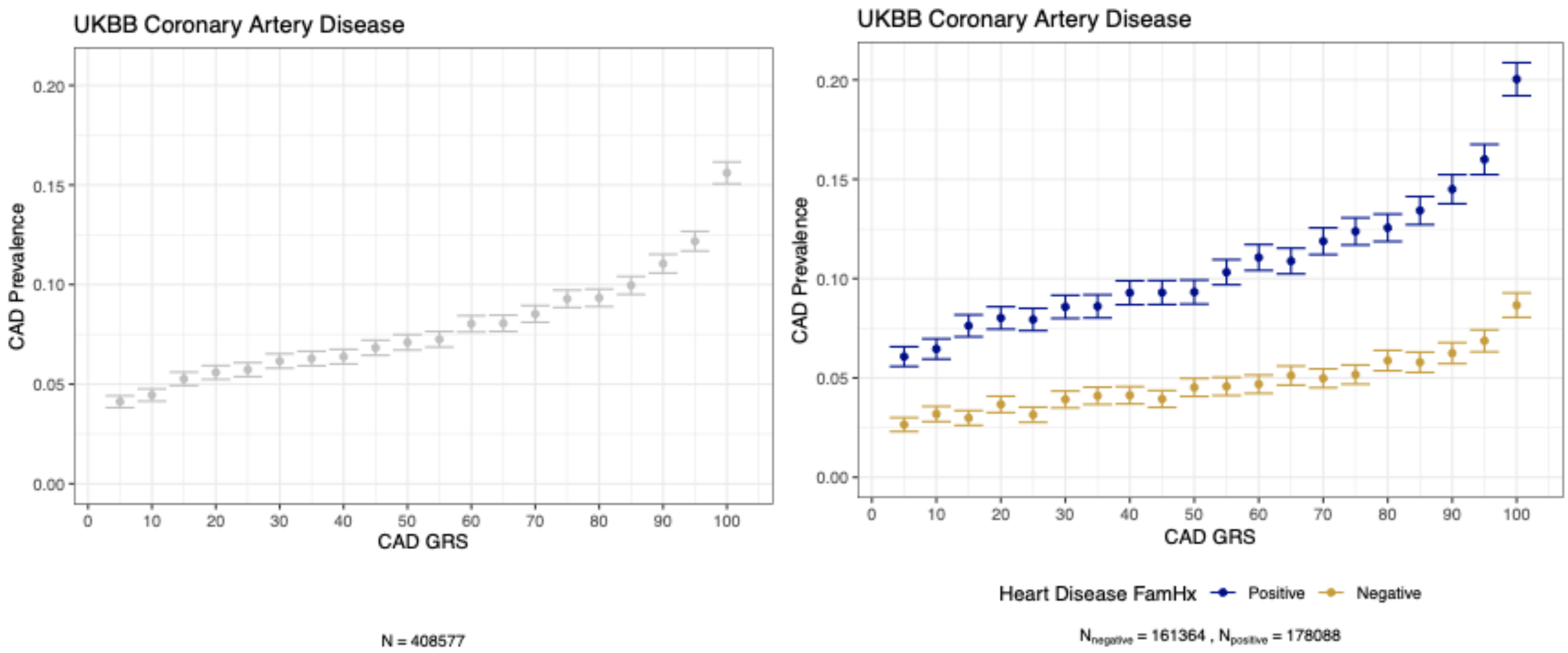


**Supplementary Figure 6 CAD prevalence in UK Biobank by PGS ventiles and family history strata**

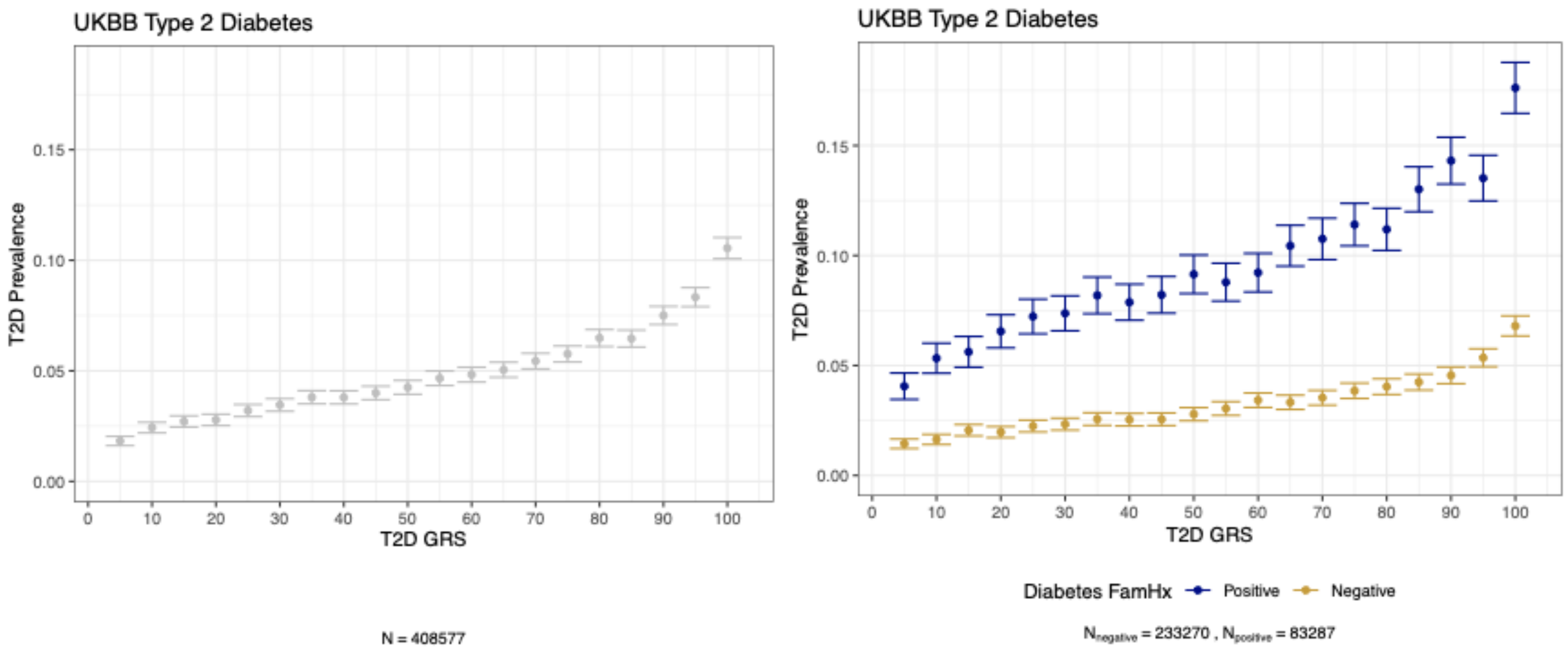


**Supplementary Figure 7 T2D prevalence in UK Biobank by PGS ventiles and family history strata**

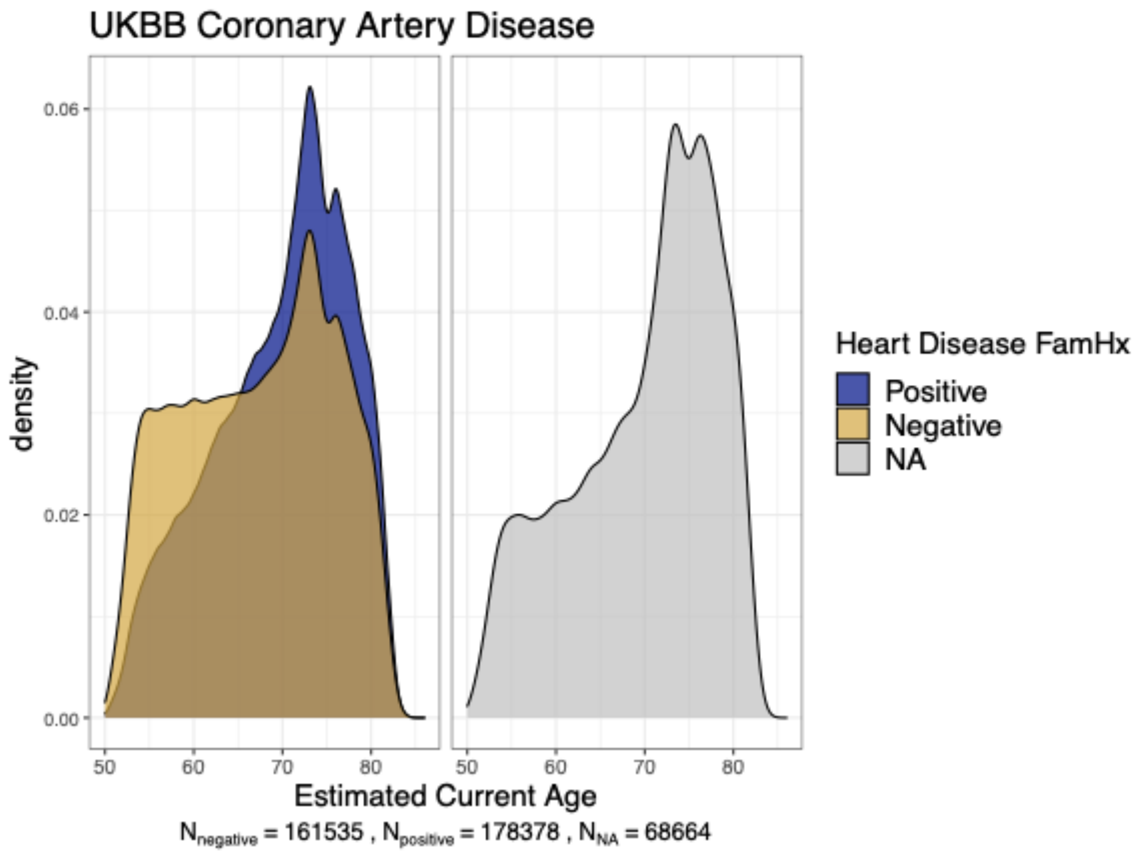


**Supplementary Figure 8 Age distribution in UK Biobank.** With recent enrollment and only one time point, we are using current age to estimate the age of self-reported family history of heart disease in UK Biobank.

**Supplementary Table 1 Clinical impact of high risk stratification for T2D in HUNT.**

| Predictor | High Risk definition | Reference Group | Odds Ratio | 95% CI | p-value | % of sample in High Risk | Median participati on age in High Risk | Prevalence in High Risk | Prevalence in Reference Group | Sensitivity | Specificity |
|---|---|---|---|---|---|---|---|---|---|---|---|
| PGS | Top 20% | Remaining 80% | 2.09 | 1.97-2.24 | $4.15x10^{-113}$ | 20 | 40.7 | 0.123 | 0.066 | 0.32 | 0.81 |
| | Top 10% | Remaining 90% | 2.82 | 2.11-2.47 | $7.83x10^{-93}$ | 10 | 40.8 | 0.119 | 0.071 | 0.18 | 0.91 |
| | Top 5% | Remaining 95% | 2.35 | 2.35-2.88 | $3.02x10^{-75}$ | 5 | 41.0 | 0.116 | 0.073 | 0.10 | 0.95 |
| | Top 1% | Remaining 99% | 2.85 | 2.31-3.52 | $1.67x10^{-22}$ | 1 | 40.9 | 0.109 | 0.077 | 0.02 | 0.99 |
| FH | Positive | Negative | 3.12 | 2.91-3.36 | $2.44x10^{-212}$ | 22.9 | 52.6 | 0.159 | 0.053 | 0.47 | 0.79 |
| PGS conditional on Positive FH | Top 20% of Positive FH | Remaining 80% | 3.14 | 2.85-3.46 | $6.21x10^{-119}$ | 4.6 | 51.5 | 0.223 | 0.071 | 0.13 | 0.96 |
| | Top 10% of Positive FH | Remaining 90% | 3.54 | 3.13-4.02 | $6.90x10^{-87}$ | 2.3 | 51.2 | 0.253 | 0.074 | 0.07 | 0.98 |
| | Top 5% of Positive FH | Remaining 95% | 3.65 | 3.07-4.32 | $6.83x10^{-50}$ | 1.1 | 51.1 | 0.265 | 0.075 | 0.04 | 0.99 |
| | Top 1% of Positive FH | Remaining 99% | 4.39 | 3.06-6.31 | $1.07x10^{-15}$ | 0.23 | 51.0 | 0.299 | 0.077 | 0.01 | 0.99 |

An indicator variable was created for the various high risk definitions above. The model controlled for batch, participation age, participation age squared, birth year, principal components 1-4 from genetic data, and sex.

**Supplementary Table 2 Full model estimates for T2D in both cohorts.**

| | HUNT | | | UKB | | |
|---|---|---|---|---|---|---|
| **Predictor** | **OR** | **95% CI** | **p-value** | **OR** | **95% CI** | **p-value** |
| Standardized Participation Age | 2.22 | 1.76-2.74 | $2.61x10^{-13}$ | 0.70 | 0.53-0.93 | 0.014 |
| Standardized Participation Age Squared | 0.46 | 0.39-0.56 | $2.54x10^{-16}$ | 0.86 | 0.66-1.11 | 0.235 |
| Standardized 2021-birthYear | 2.22 | 2.06-2.40 | $1.80x10^{-97}$ | 2.83 | 2.64-3.03 | $3.72x10^{-192}$ |
| Male Sex | 1.41 | 1.33-1.50 | $2.18x10^{-30}$ | 1.96 | 1.90-203 | $< 2.2 \times 10^{-308}$ |
| Positive Family History | 3.01 | 2.79-3.24 | $5.58x10^{-181}$ | 3.01 | 2.90-3.11 | $< 2.2 \times 10^{-308}$ |
| Inverse normalized PGS | 1.60 | 1.54-1.67 | $9.65x10^{-115}$ | 1.52 | 1.49-1.56 | $1.56x10^{-265}$ |
| Family History x Inverse normalized PGS | 0.913 | 0.86-0.97 | 0.0032 | 0.99 | 0.95-1.02 | 0.42 |

Adjusted for principal components 1-4 from genetic data and genotyping batch (HUNT)/genotyping array (UKB).

**Supplementary Table 3 Model comparisons for T2D in HUNT**

| Model 1 | Model 2 | LRT p-value | Nagelkerke's $r^2$ |
|---|---|---|---|
| Base | PGS model | $2.55x10^{-202}$ | 0.031 |
| Base | FH model | $5.95x19^{-214}$ | 0.033 |
| PGS model | PGS + FH (additive) model | $6.84x10^{-185}$ | 0.028 |
| FH model | PGS + FH (additive) model | $2.94x10^{-173}$ | 0.026 |
| PGS + FH (additive) model | PGS + FH + PGS x FH (interaction) model | 0.003 | 0.00029 |

**Supplementary Table 4 Full model estimates for CAD in UKB**

| Predictor | OR | 95% CI | p-value |
|---|---|---|---|
| Standardized Participation Age | 1.35 | 1.054-1.74 | 0.0179 |
| Standardized Participation Age Squared | 0.54 | 0.43-0.67 | $3.6 \times 10^{-8}$ |
| Standardized 2021-birthYear | 3.02 | 2.87-3.19 | $< 2.2 \times 10^{-308}$ |
| Male Sex | 2.87 | 2.79-2.95 | $< 2.2 \times 10^{-308}$ |
| Positive Family History | 2.03 | 1.98-2.1 | $< 2.2 \times 10^{-308}$ |
| Inverse normalized PGS | 1.41 | 1.38-1.44 | $1.29 \times 10^{-169}$ |
| Family History x Inverse normalized PGS (interaction term) | 1.03 | 1.01-1.07 | 0.0134 |

Adjusted for principal components 1-4 from genetic data and genotyping array.

**Supplementary Table 5 Model comparisons for CAD and T2D in UKB**

| | | CAD | | T2D | |
|---|---|---|---|---|---|
| **Model 1** | **Model 2** | **LRT p-value** | **Nagelkerke's $r^2$** | **LRT p-value** | **Nagelkerke's $r^2$** |
| Base | PGSmodel | $< 2.2 \times 10^{-308}$ | 0.0234 | $< 2.2 \times 10^{-308}$ | 0.296 |
| Base | FH model | $< 2.2 \times 10^{-308}$ | 0.0207 | $< 2.2 \times 10^{-308}$ | 0.046 |
| PGS model | PGS + FH (additive) model | $< 2.2 \times 10^{-308}$ | 0.0168 | $< 2.2 \times 10^{-308}$ | 0.0378 |
| FH model | PGS + FH (additive) model | $< 2.2 \times 10^{-308}$ | 0.0195 | $< 2.2 \times 10^{-308}$ | 0.0218 |
| PGS + FH (additive) model | PGS + FH + PGS x FH (interaction) model | 0.014 | 0.00004 | 0.416 | $6.2 \times 10^{-6}$ |

.

**Supplementary Table 6. Cohort demographics**

| | **HUNT Study (N=69,635)** | **UK Biobank (N=408,577)** |
|---|---|---|
| Median participation age range for family history (IQR) | 41.4 (30.4,54.9) | 59 (51,64) |
| Median birth year (IQR) | 1951 (1937,1964) | 1950 (1945,1957) |
| Female (%) | 53 | 54 |
| T2D prevalence (%) | 7.8 | 4.9 |
| CAD prevalence (%) | 9.7 | 7.8 |
| MI prevalence (%) | 9.0 | NA |

**Supplementary Table 7 Phenotype definitions for main outcomes and family history variables in HUNT and UKB**

| | **HUNT** | | **UKB** | |
|---|---|---|---|---|
| | **CAD** | **T2D** | **CAD** | **T2D** |
| **Case definition** | Self reported CABG or PCI or MI ICD code (I21,I25.2,410,412) | Non fasting serum glucose > 11.1, HbA1C > 6.5 or E11, 250.00, 250.02, 250.10, 250.12, 250.20, 250.2, 250.30, 250.32, 250.40, 250.42, 250.50, 250.52, 250.60, 250.62, 250.70, 250.72, 250.80, 250.82, 250.90, 250.9 | Phecode 411 for ischemic heart disease | Phecode 250.2 for Type 2 diabetes |
| **Self-reported family history** | HUNT1: Sibling with heart attack or angina pectoris<br>HUNT2: Parents or siblings had an MI or chest pain AND Mother, Father, Sister, Brother, Child had heart attack before age 60<br>HUNT3: Parents, siblings or children had heart attack before age 60 | HUNT1: Siblings with diabetes<br>HUNT2: Mother, father, brother, sister, child with diabetes<br>HUNT3: Parents, siblings or children with diabetes | Heart disease of mother, father, or sibling | Diabetes of mother, father, or sibling |

**Supplementary Table 8 Sample sizes.**

| | Self-reported family history | Control | Case | Total |
|---|---|---|---|---|
| HUNT CAD | Negative | 41,361 | 2,923 | 44,284 |
| | Positive | 20,705 | 3,741 | 24,446 |
| | Unknown/NA | 827 | 78 | 905 |
| HUNT T2D | Negative | 48,886 | 2,760 | 51,646 |
| | Positive | 12,926 | 2,445 | 15,371 |
| | Unknown/NA | 2,441 | 177 | 2,618 |
| UKBB CAD | Positive | 159,012 | 19,076 | 178,088 |
| | Negative | 153,762 | 7,602 | 161,364 |
| | NA | 63,143 | 5,387 | 68,530 |
| UKBB T2D | Negative | 225,772 | 7,498 | 233,270 |
| | Positive | 57,380 | 7,907 | 65,287 |
| | Unknown/NA | 87,217 | 4,790 | 92,007 |

The number of cases and controls and self-reported positive/negative family history participants in UKB and HUNT for both CAD and T2D.
